# Consumption of breast milk, formula and other non-human milk by children aged under two years: analysis of 86 low and middle income countries

**DOI:** 10.1101/2020.06.02.20120519

**Authors:** Paulo A. R. Neves, Aluisio J.D. Barros, Phillip Baker, Ellen Piwoz, Thiago M. Santos, Giovanna Gatica-Domínguez, Juliana S. Vaz, Nigel Rollins, Cesar G. Victora

**Affiliations:** International Center for Equity in Health, Universidade Federal de Pelotas. Rua Marechal Deodoro, 1160, 3^rd^ floor. 96020-220, Pelotas, Brazil.; Institute for Physical Activity and Nutrition, Deakin University. 221 Burwood Highway, VIC 3125, Melbourne, Australia.; Global Development Program, The Bill & Melinda Gates Foundation.PO Box 23350, 98102-0650 Seattle, WA, United States.; Department of Maternal, Newborn, Child and Adolescent Health, World Health Organization.Avenue Appia 20, CH-1211, Geneva 27, Switzerland.

**Author notes:** Corresponding author:* Paulo A. R. Neves Rua Marechal Deodoro, 1160, 3^rd^ floor, 96020-220, Pelotas, Brazil.

## Abstract

**Background:** Consumption of breast milk substitutes (BMS) by children aged under six months in low and middle income countries (LMICs) is directly driven by country income and family wealth. Multi-country investigations on the consumption of BMS by older children (6–23 months) are lacking.

**Methods:** Using data from 86 nationally representative surveys carried out in LMICs from 2010 onwards, we analyzed the prevalence of continued breastfeeding at one and two years, and frequency of consumption of formula and other non-human milk by age in months. Indicators were estimated through 24-hour dietary recall. Absolute and relative wealth indicators were used to describe within- and between-country socioeconomic inequalities. Results were stratified by country income groups.

**Findings:** Breastfeeding declined sharply as children became older in all LMICs, especially in upper-middle income countries. Formula consumption peaked at six months of age in low and lower-middle income countries, and at around 12 months in upper-middle income countries. Consumption of formula at any age higher in children from wealthier families in all countries, while breastfeeding was more common among poor children. Multilevel linear regression analysis showed that consumption of formula was positively associated while breastfeeding was negatively associated with absolute national income. Factors at country level explained a substantial proportion of overall variability in formula use and breastfeeding.

**Interpretation:** Infant and young child feeding practices vary strongly according to wealth, both within and between countries. Breastfeeding falls sharply as children become older, especially in wealthier families living in upper-middle income countries; this is also the group with highest formula consumption at any age. Country-level factors play an important role in explaining BMS consumption by all family wealth groups in LMICs, suggesting that formula marketing at national level may be partly responsible for the observed differences.

**Funding:** The Bill & Melinda Gates Foundation, through the WHO

## Introduction

Breastfeeding is essential for children, their mothers, and societies as it is associated with reduced risk of short- and long-term harmful health outcomes in all country contexts.^1,2^ Breastfeeding also associates with increases in intelligence and educational attainment,^3^ hence contributing to workforce productivity and sustainable development.^2,3^ Despite the recommendation by the WHO for all children to be breastfed to 24 months or longer, the prevalence of continued breastfeeding at one year remains below 60% in upper middle income countries and under 30% in high income countries,^1^ being inversely correlated with national wealth.^1,4^

The types of milk consumed by young children include three broad categories: breastmilk, commercial breastmilk substitutes (BMS) or other types of non-human milk. One of the major threats to achieving optimal breastfeeding practices in all countries is the aggressive marketing of BMS, including formula.^2,4^ Recent studies have shown an escalation in BMS sales worldwide, mostly concentrated in high and upper middle income countries,^5,6^ and for all BMS categories: standard/infant formula (for children aged 0–6 months), follow-up (6–12 months) and toddler (13–36 months) formula. This is despite the WHO having long considered products in these latter categories as unnecessary and unsuitable as BMS.^7^ Recent evidence from the United States suggests that toddler milk sales has grown rapidly, with annual sales volumes growing by 158% for toddler milk and decreasing by 7% for infant formula between 2006–2015.^6^ Similar trends have been reported for emerging economies with rapid increases in per capita sales volumes of follow-on and toddler milk, especially in China, Brazil, Peru, and Turkey.^5^

Between- and within-country socioeconomic inequalities in the consumption of BMS by infants in low- and middle-income countries (LMICs) have been reported previously, showing that the consumption of formula is directly correlated with national wealth.^4^ Major gaps between rich and poor families were seen in some regions of the world, such as in Latin America & the Caribbean and in East Asia & the Pacific. Additionally, infant formula consumption was inversely correlated with the prevalence of breastfeeding at one year at national level. In addition, breastfeeding was more common among poor than rich children in LMICs.^4^

As the per capita volume sales of all formula categories is rapidly escalating in LMICs – and particularly in upper middle income countries^5^ – it is important to document the frequency of BMS consumption. Earlier analyses focused on children aged below six months,^4^ but given promotion of formula for older children^5,6^ it is essential to document consumption among children aged 6–23 months.

This study aimed at investigating the prevalence and socioeconomic inequalities in the consumption of types of milk by children aged 6–23 months in LMICs. We give special attention to socioeconomic inequalities in infant and young child feeding (IYCF) practices, exploring the relationship between the consumption of different types of milk and relative and absolute wealth levels.

We hypothesized that: a) as children become older, breastfeeding and consumption of formula decline, while consumption of other types of non-human milk increases; b) as national wealth increases, consumption of formula increases and breastfeeding decreases; and c) as formula is an expensive product for most LMICs families, its acquisition might be directly associated with a certain absolute level of income.

## Methods

For the present investigation, we relied upon nationally representative cross-sectional surveys conducted periodically in LMICs, namely Demographic Health Surveys (DHS)^8^ and Multiple Indicator Cluster Surveys (MICS).^9^ Such surveys cover a large number of reproductive, maternal, newborn, and child health indicators, employing multistage sampling strategies to collect data at household level. We selected the most recent publicly available survey for each country carried out since 2010. In both types of surveys, face-to-face interviews were performed through standardized questionnaires, so that both types of surveys are highly comparable.^10^ Women of childbearing age (15–49 years) were interviewed by trained fieldworkers, who collected data on IYCF practices using 24-hour recall for the youngest child born in the last two years before the survey.^11^ We also included the nationally representative surveys conducted in Ecuador and Peru, after harmonization of its dataset and variables in accordance with the DHS/MICS standards.^12,13^

Data for more than 100 surveys were available at the time of the analysis. However, some surveys had to be excluded, as detailed in Supplementary figure 1. Therefore, our analysis are based on data for 86 LMICs.

We calculated the following feeding indicators: *continued* breastfeeding *at one and two years* (proportion of children between 12–15 and 20–23 months of age, respectively, who were breastfed);^11^ *formula consumption under six and between 6–23 months* (proportion of children between 0–5 months and 6–23 months who were fed formula); and *other non-human milk consumption under 6 and between 6–23 months* (proportion of children between 0–5 months and 6–23 months who were fed non-human milk, other than formula, e.g. cow and goat milk). The consistency and quality of the calculated estimates was checked by comparison with the published figures in the DHS/MICS reports for continued breastfeeding at one and two years;^11^ all differences were < 1 percentage point. In our analyses, we use the terms formula and BMS as synonyms.

Countries were grouped according to World Bank income classification at the year of survey implementation.^14^ The supplementary table 1 provides a list of countries included in the analyses, their classification, and the sample size by age range. All regional and income estimates were weighted by the population size of children in each age-range in the country retrieved from the World Bank Population Estimates and Projections, in the year the survey was carried out.^15^

To analyze within-country socioeconomic inequalities we used the wealth index provided with each survey dataset, based on assets (television, refrigerator, car, etc.) and building characteristics (wood floor, brick wall, and roof). The index was derived using principal component analysis.^16^ Two separate analyses were carried for urban and rural households, later combined into a single score using a scaling procedure to allow comparability between areas of residence. The definition of area of residence is country-specific. The resulting index was then split into five groups of equal sample size – the quintiles. The first quintile represents the households with the poorest 20% of the population, and the fifth quintile the wealthiest 20% of the sample.^17^

Furthermore, we attributed absolute income values to each wealth quintile based on Fink et al.^18^ methods. Briefly, the absolute income is estimated for each wealth quintile based on the national income levels (gross domestic product) retrieved from the World Bank, and the national income inequality data obtained from the Standardized World Income Inequality Database Gini index, to generate parameters of a log-normal distribution.^19,20^ Considering the household asset index, dollar values are then assigned to each wealth quintile. Absolute income is expressed in 2011 purchasing power parity adjusted international dollars.

We estimated consumption of each type of milk by age in months, and then used local polynomial smoothing to graph trajectories of consumption of the three types of milk by age for countries and for the poorest and richest quintiles in each country. We also explored the share of formula consumption among all types of non-human milk (formula plus other nonhuman milk) consumed by children aged under two years (supplementary table 2).

All estimates were calculated using the svy command in Stata. Multilevel linear regression models were performed to explore the relationship between feeding indicators and absolute income (log-transformed to better fit the models). Countries were deemed the highest hierarchical level, and quintiles within each country as the second hierarchical level. Analysis were adjusted by the World Bank income classification, and interaction terms were fitted. We present beta coefficients (in percent points) alongside with 95% confidence intervals (CI) for each outcome, as well as the proportion of the overall variance explained by the highest level (country). We graphed scatter plots to illustrate the relation between feeding indicators and absolute income, including predicted lines for each income level group. Fractional polynomials were used to describe non-linear associations in graphical form. In the multilevel analyses, however, using polynomials did not improve the fit of the model compared to a linear equation, and for the sake of simplicity we adopted the linear regression. Lastly, we plotted within-country inequalities for the top three and bottom three countries in terms of the consumption of formula and of breastmilk. All analyses were run using Stata 16·0 (Stata Corp.) and the graphs built up on R (version 3·6·1).

This study is based on anonymous publicly available datasets, so any ethical clearance was the responsibility of the national institutes in charge of data collection. National estimates and by wealth quintiles are given for each indicator with the 95% CI (Supplementary tables 3–8).

## Results

Data were available from 86 countries (Supplementary figure 1), with sample sizes ranging from 635 children aged under two years in Kosovo to 94,371 in India. The median year of the surveys was 2014, ranging from 2010 (Bhutan, Burkina Faso, Central African Republic, Colombia, and South Sudan) to 2018 (Iraq, Kyrgyzstan, Peru, Suriname, and Tunisia). Studied countries represented 41·6% of upper middle, 70·2% of lower middle and 90·3% of low income countries.

Figure 1 shows the proportions of children receiving breastmilk, formula and other types of milk, by age and country income groups. Breastfeeding prevalence falls with increasing age in all income groups below the global recommendation for continued breastfeeding beyond two years, particularly in upper-middle income countries. Formula use is most common in middle income groups. It increases slightly with age in all country income groups, but after 6–9 months it tends to decline. Other milk consumption increases with age during the first year of life, and remains stable thereafter. In low income countries, formula and other types of milk are used by similar proportions of children at birth, but formula consumption remains stable whereas other milks increase in frequency. A higher proportion of children consumed formula than other types of milk during the first five months in lower middle income countries, and during the first 10–11 months in upper middle income countries. In all groups of countries, other milks are more frequently used than formula and less than half of children were breastfed at 24 months of age.

**Figure 1.**
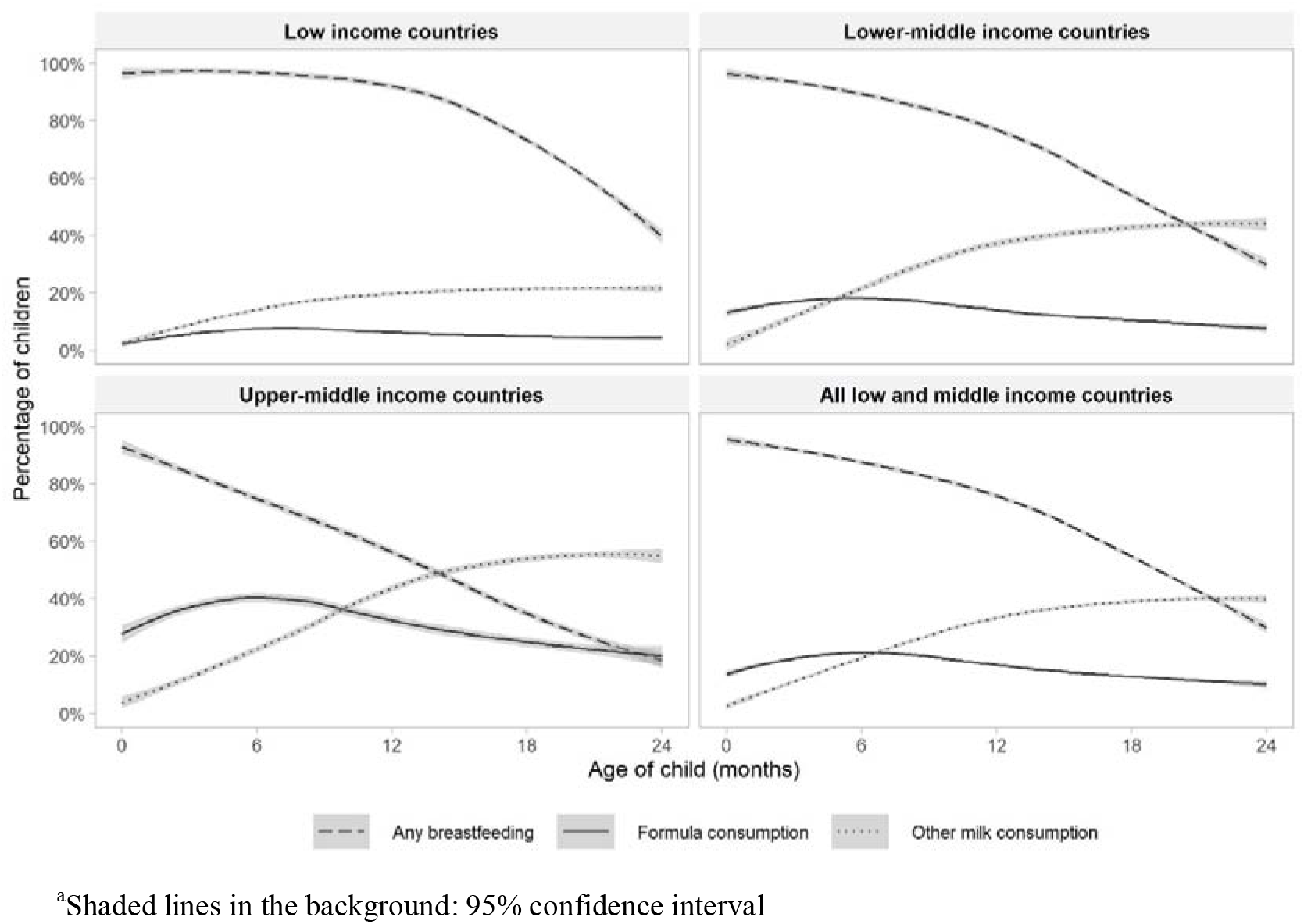
Trajectories of the frequency of consumption of different types of milk by children’s age^a^.

Figure 2 expands upon the results in Figure 1, showing feeding patterns for the wealthiest and poorest quintiles in each group of countries. In all groups, breastfeeding is less common, and consumption of non-human milk is more common among children from the richest families, especially for formula consumption. The exception occurs in upper middle income countries, where consumption of other types of milk is almost equally prevalent for children from the different quintiles. The only group where formula becomes more common than breastfeeding are children from the wealthiest quintile in upper middle income countries, where this occurs at around 20 months of age.

**Figure 2.**
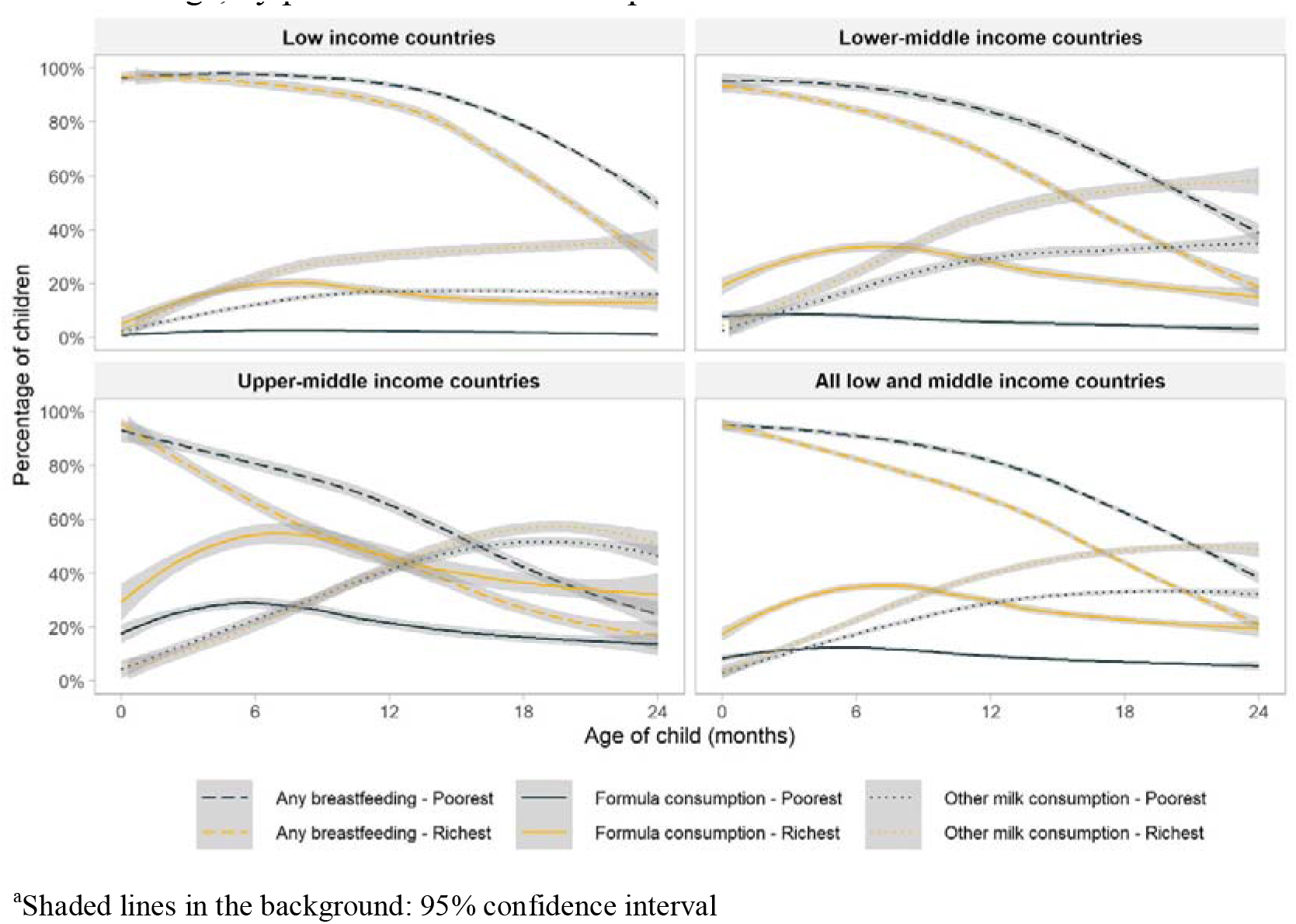
Trajectories of the frequency of consumption of different types of milk by children’s age, by poorest and wealthiest quintiles^a^.

Based on the fitted curves shown in Figures 1 and 2, formula consumed by children as a proportion of all types of non-human milk combined was 69%, 35%, 26%, and 17% at 0, 6, 12, and 24 months of age, respectively, in low income countries; for the same age groups, in lower middle income countries the shares were 94%, 45%, 29%, and 16%, and in upper middle income countries 97%, 63%, 44%, and 28%. When the analyses are broken down by wealth quintiles, the results confirm the higher use of formula among children from wealthier families, compared to those in poor families, in all country income groups (Supplementary table 2).

Figure 3 illustrates the inverse, non-linear relationship between breastfeeding at one and two years and absolute income across all age ranges and income groups, although the pattern is less marked for continued breastfeeding at two years in upper middle income countries. Findings for formula use are almost a mirror-image of those for breastfeeding. In all age-ranges, consumption of formula increases with higher absolute income. At a higher absolute income level, use of formula escalates in all groups of countries.

**Figure 3.**
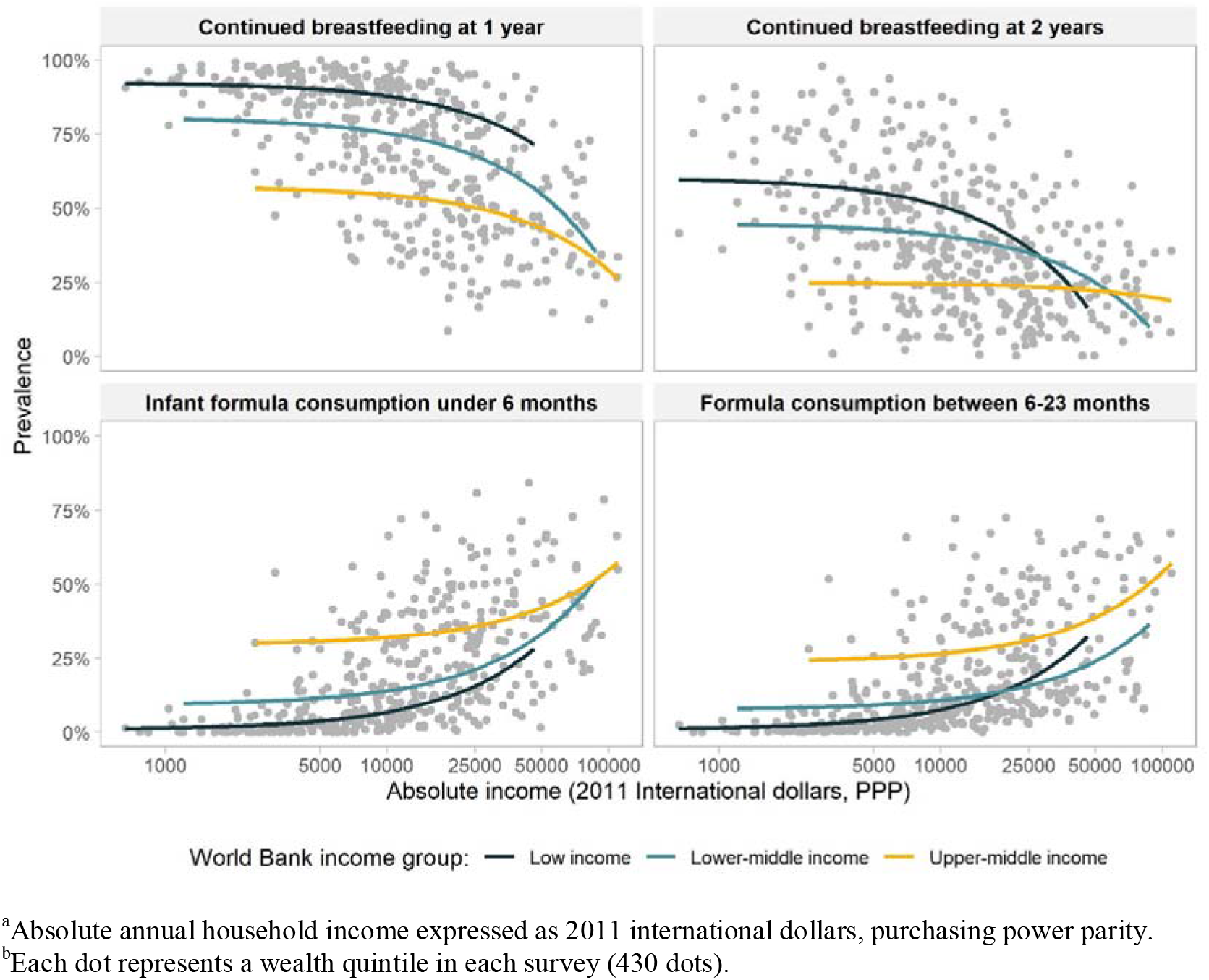
Relationship between feeding practices of children and absolute income^a,b^.

Patterns for other types of milk are less straightforward (Supplementary figure 2). Among young infants, the association with household income is weak; however, for children aged 6–23 months, the use of other milks becomes more common with increasing household income, particularly in low and lower middle income countries. As observed for formula and breastmilk, there are large differences between countries for high or low consumption of other milks, at the same level of absolute income.

A common finding in the four charts included in Figure 3 is that for the same level of absolute household income, breastfeeding rates tend to be higher in low income than in middle income countries, while the reverse pattern is observed for formula. This suggests that characteristics of the countries other than income alone may be driving feeding patterns. Figure 4 is an expanded version of Figure 3, showing the top three and the bottom three countries according to national prevalence of use of each type of milk. All countries in the top formula consumption groups belong to the upper middle income category, whereas those in the low consumption group are low income countries. For breastfeeding, all quintiles in the high-consumption group of countries show substantially higher rates, regardless of household income levels. The opposite pattern is observed for formula, where at similar household income levels there are striking differences between the three top and bottom consumer countries. Even the poorest households in the top consumers countries used more formula than the wealthiest quintiles in bottom consumers countries.

**Figure 4.**
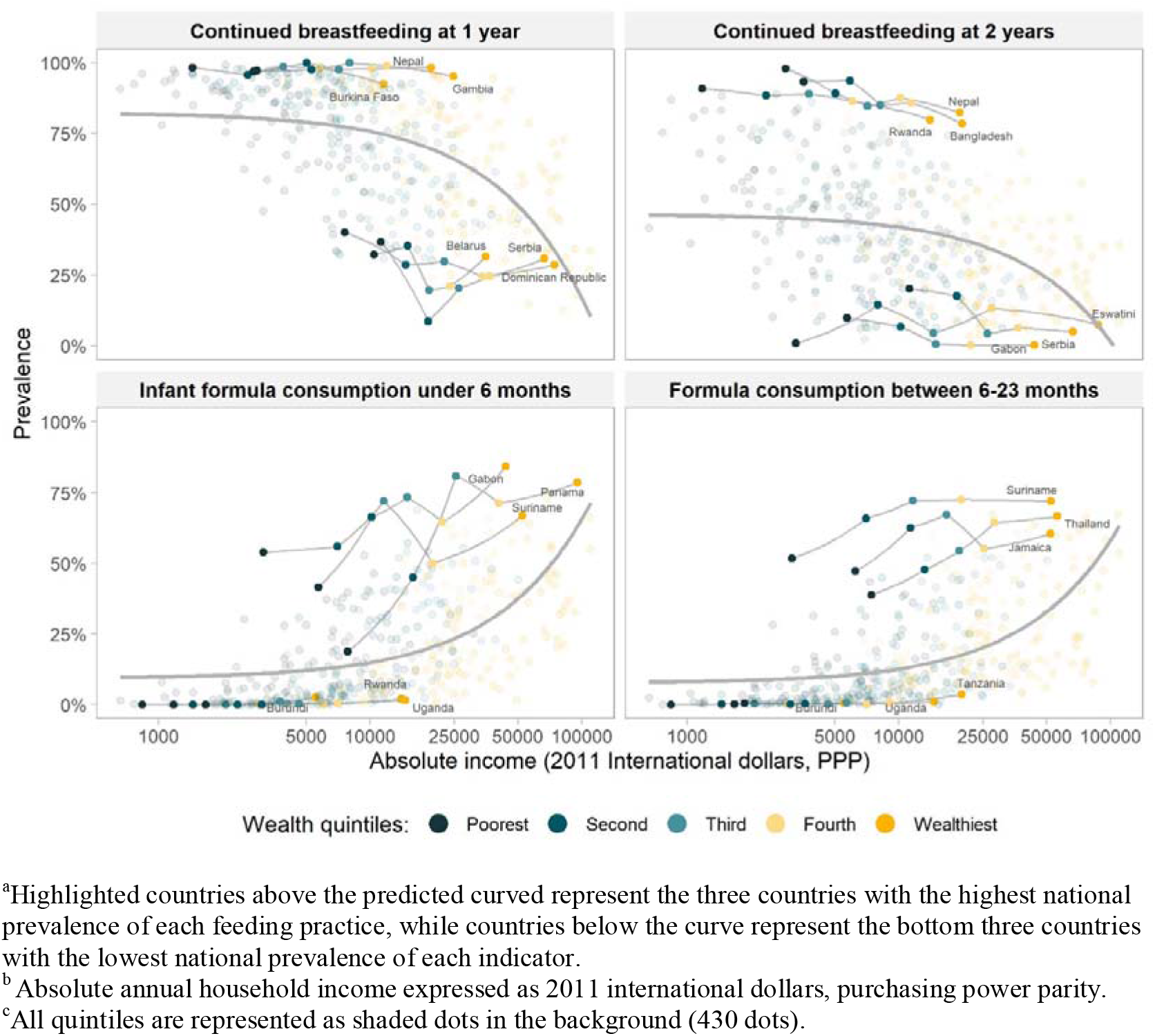
Top and bottom countries^a^ in relation to the feeding practices and absolute income^b^ and relative wealth^c^.

We further explored the heterogeneity among countries using multilevel analyses, with countries representing the first level and log household income per quintile within each country in the second level (Table 1). In this type of analysis, it is common to observe that the higher level variable –countries – explains a small proportion of the overall variance. Yet in this analysis, we found that this level explained nearly 30% of the variance in infant formula consumption under six months and of continued breastfeeding at one year, thus suggesting the existence of country characteristics that markedly influence feeding patterns by all groups of wealth, which deserve more in-depth analysis in the future.

**Table 1.** Multilevel association between feeding indicators and log absolute income^a^, adjusted by World Bank income level.

| Indicator | Unadjusted $\beta$<br>(95% CI)<sup>b</sup> | Adjusted $\beta$<br>(95% CI)<sup>b</sup> | Variance<br>(%)<sup>c</sup> | P |
| --- | --- | --- | --- | --- |
| Continued breastfeeding at 1 year | -8.0 (-9.0; -7.0) | -9.0 (-10.8; -7.2) | 27.3 | <0.0001 |
| Continued breastfeeding at 2 years | -8.2 (-9.3; -7.1) | -3.4 (-5.3; -1.5) | 21.2 | <0.0001 |
| Infant formula consumption under 6 months | 8.1 (7.2; 9.0) | 8.5 (6.8; 10.1) | 29.4 | <0.0001 |
| Formula consumption between 6-23 months | 8.7 (8.0; 9.4) | 10.6 (9.4; 11.8) | 17.6 | <0.0001 |
| Other milk consumption under 6 months | 1.0 (0.4; 1.5) | -0.7 (-0.1; 0.3) | 20.0 | 0.171 |
| Other milk consumption between 6-23 months | 6.1 (5.2; 7.0) | 2.2 (0.7; 3.7) | 15.4 | 0.004 |

<sup>a</sup>Absolute annual household income expressed as 2011 international dollars, purchasing power parity
<sup>b</sup>Betas represent changes in each indicator as percentage points by increments of 1 log of absolute income.
<sup>c</sup>Variance percentage that is explained by the highest hierarchical level in the adjusted analysis: countries.

## Discussion

This is the first investigation to describe multi-country use of formula and other types of nonhuman milk among children aged 6–23 months. We confirm the well-known finding that breastmilk consumption decreases with the age of the child, across all country income groups, and that early cessation of breastfeeding is more common among children from wealthy families in all groups of countries.^1,21^ Patterns for formula use peak at around 6–9 months of age in low and lower middle income countries, and somewhat later (up to 12 months) in upper middle income countries. Formula use in the second year of life is relatively common in children from wealthier families in upper middle income countries where it accounts for about 40% of all children until this age. At the other extreme, formula is used by less than 10% of children from the poorest families in low income countries, throughout the age range from birth to two years.

Our findings further show that other types of non-human milk are an important feeding source for children under two years in all LMICs, especially in middle income countries, and that formula is progressively replaced by these types of milk as children become older. However, this varies markedly by family wealth and country income group. These foods are rarely explored in the IYCF literature with most available studies reporting on associations with health outcomes, such as allergy risk or child growth.^22,23^

These results add further evidence that follow-on and toddler milks are now prominent in the diets of children worldwide, and especially in children living in middle income countries.^5,24^ Guidance from WHO deems follow-up and toddler formula unnecessary and unsuitable as BMS, and recognizes these products have partially or totally replaced the consumption of breastmilk.^7,25^ Following this guidance, in 2016 the World Health Assembly adopted a new resolution clarifying that BMS, as defined by the International Code of Marketing of Breastmilk Substitutes (CODE), includes any milk products marketed for feeding infants and children aged 0–36 months.^25^ As of 2018, however, only 44 countries restricted the marketing of BMS beyond 12 months of age, and just 16% of countries with existing CODE provisions in place cover products for 0–36 months.^7^

Subsequently, the marketing of BMS for children beyond 12 months of age is not legally restricted in the large majority of countries. In the United States, for example, advertising expenditure on toddler milks increased 4-fold between 2006–2015 and sales volume increased 2·6-fold, while advertising expenditure and volumes of infant formula declined.^6^ Another reason for concern is that the branding, packaging and labelling of these products frequently resembles and is often mistaken by parents and caregivers for infant formula.^26^ Companies use this ‘cross-promotion’ strategy to boost brand loyalty across their entire BMS product range, make actual or implied product claims, and indirectly promote infant formula in countries where legislation prohibits this.^25,27^ The importance of national level policies and marketing regulations is reinforced by our finding in the multi-level analyses showing a substantial proportion of the variation in formula use is explained at the country level; in particular, consumption by children in households with the same level of absolute wealth varies markedly depending to the country were the children live.

As early introduction of formula (under six months) hampers optimal breastfeeding practices,^4^ formula advertisement for infants and older children must end promptly. There is a growing body of evidence demonstrating that marketing of formula, through health facilities, in mass and digital media, and via labeling and point of sale promotion undermines optimal breastfeeding practices wolrdwide.^5,28^ Additionally, promotion of BMS among health professionals and facilities negatively affects breastfeeding practices, as women tend to follow practitioners advise and replace the breast milk by BMS.^26^ This marketing occurs in countries despite national adoption of the CODE, being its enforcement urgently needed. In LMICs, however, other non-human milk depicts an important share among the feeding sources to children after six months of age, in general.

Noteworthy is the fall of continued breastfeeding in all income groups in LMICs as children become older. As earlier breastfeeding ceases, there are potentially implications for growth and development of children associated with early introduction of unhealthy foods, sub-optimal dietary diversity and lower consumption of nutrient-rich foods.^29^

The main drivers of breastfeeding, and conversely of formula feeding, were summarized by Rollins et al.^2^ These include social trends; media and promotion of products in health systems and the community; support in health systems, family, community, and in the work place; and individual-level factors, such as mother attributes and relationship between mother-child.^2^ A complex relationship is described between breastfeeding, formula use, women’s work, and income^2^ that our analysis also corroborates. Over the past decades, more women have entered the formal work force, which affects breastfeeding patterns globally. While paid employment may make formula feeding more affordable, and formula options may allow women to return to work earlier after childbirth, there are nonetheless millions of working women who want to breastfeed optimally but are unable to because of maternity leave policies and working environments are not supportive. Part of this can be attributed to the miss commitment and lack of obligation by countries in adopting comprehensively the implementation of the CODE, that protects women and children against aggressive BMS marketing and advertisings, enabling women to breastfeed longer.^2,7,26^

The strengths of our analyses include the large number of countries studied, high comparability in methods and field procedures in all surveys, reliance on feeding data collected through 24-hour recall (thus reducing recall bias) and the use of absolute income, given the high cost of formula around the world, especially for most families in LMICs. Absolute income has also been shown to predict other maternal and child health outcomes.^18,30^ Some limitations are acknowledged, as the lack of data for high income and emerging economies countries that represent large markets for formula companies; the exclusion of six surveys (out of the 100 eligible) due to small sample sizes in each wealth quintile, and the lack of information to calculate absolute income for three surveys.

## Conclusion

Globally, there is growing interest in so-called “commercial determinants of health”, represented by unhealthy promotion of tobacco, alcohol and ultra-processed foods, among others. BMS promotion clearly constitutes one type of such determinants.^31^ Our analyses of breastmilk and BMS consumption confirmed that wealth is an important determinant of IYCF practices. However, we also show that there is wide variability in consumption at the same level of family income across countries, and that country level variables play an important role. This suggests that formula marketing at country level leads to greater use by all socioeconomic groups within a country. Further analyses are required to explore the country level determinants of such variability.

## Data Availability

All data can be obtained accessing the websites of each organization in charge of data collection and disclosure.

https://dhsprogram.com/

http://mics.unicef.org/

## Author’s contribution

PARN carried out the analysis, with technical support from AJDB and GGD. TMS contributed with graphic design. PARN and CGV interpreted the results and wrote the manuscript. PB, EP, JSV, and NR contributed with the discussion. All authors read and approved the final version of the manuscript.

## Acknowledgments

We are thankful to Cauane Blumenberg and Luiz Paulo V Ruas for the statistical insights generously provided. This work was funded by The Bill and Melinda Gates Foundation, through the World Health Organization (grant numbers: OPP1179886/ INV-007594) and the Associação Brasileira de Saúde Coletiva. PARN was supported by the Brazilian National Council for Scientific and Technological Development – CNPq (grant number: 155541/2018–8). The views expressed are of the authors alone and do not necessarily reflect the views or policies of their respective institutions or organizations.

